# Effect of Vitamin D and Calcium Supplementation on the Kidney Function and Quality of Life of Patients with Chronic Kidney Disease (CKD): A Systematic Review and Meta-Analysis

**DOI:** 10.64898/2025.12.05.25341730

**Authors:** Hannah Guy, Abolanle Adesope, Habeeb Owolabi, Faatihah Niyi-Odumosu

## Abstract

**Objective:** This systematic review evaluates whether vitamin D and calcium supplementation benefits Chronic kidney disease (CKD) patient’s Quality of Life (QoL) and Renal function.

**Background:** CKD disrupts calcium-phosphate homeostasis, leading to secondary hyperparathyroidism and bone mineral disorders. Efficacy of vitamin D and calcium supplementation in managing CKD remains controversial.

**Methods:** PubMed, Google Scholar, Science Direct and EBSCO were searched from October 2024 to February 2025. Fifteen out of 82 identified studies were included. Data quality was assessed using Critical Appraisal Skills Programme (CASP) while bias risk was visualized using Robvis. Estimated Glomerular Filtration Rate (eGFR) and Parathyroid Hormone (PTH) outcomes were analysed using meta-analysis, whereas Bone Mineral Density (BMD) and Quality of Life (QoL) were assessed through narrative synthesis.

**Results:** Vitamin D supplementation had no significant effect on eGFR (MD=0.84mL/min/1.73m²,95%CI: –0.47 to 2.15, p=0.207) or PTH levels (MD = –6.76,95%CI: –34.92 to 1.95, p=0.005). No significant differences were found between active and inactive vitamin D supplementation on kidney function, while the results for BMD and QoL were inconclusive.

**Conclusion:** Vitamin D supplementation had no significant effects on kidney function or QoL. Limited data precluded conclusions about calcium supplementation. Future studies are needed to clarify the roles of these treatments on CKD.

**Trial Registration Number:** PROSPERO registration CRD42024607320.

- **What is already known on this topic** – Chronic Kidney Disease is a known progressive condition that affects over 10% of the global population. One of the most significant complications of CKD is the disruption if the calcium and phosphate balance, leading to conditions like secondary hyperparathyroidism. These complications can cause a significant impairment on the QoL of CKD patients. Vitamin D supplementation has become a standardised therapy for management of these complications, however there is a lack of clinical evidence in the effectiveness of these treatments.
- **What this study adds** – Despite the fact that vitamin D supplements are widely recommended to CKD patients, the evidence from the studies included in this systematic review showed no statistically significant effects. Furthermore, this review highlights the limited research on the effects of calcium supplementation, specifically in combination with vitamin D supplementation.
- **How this study might affect research, practice or policy** – This systematic review highlights the need for more rigorous clinical trials to assess the safety and efficacy of vitamin D and vitamin C supplementation in CKD patients. This research should also explore the long term effects and determine the impact of these treatments at different stages of CKD to determine whether these treatments are more effective at certain stages of the disease.

## Introduction

Chronic kidney disease (CKD) is a progressive decline in renal function, often resulting in end-stage kidney disease (ESKD), and often associated with an increased risk of comorbidities such as cardiovascular disease. These complications contribute to the acceleration of CKD progression and a reduced QoL in CKD patients^1^. CKD is clinically defined by a persistent estimated glomerular filtration rate (eGFR) of less than 60 mL/min per 1.73 m^2^ for more than 3 months, indicating impaired renal function^2^. As the eGFR levels deplete, the severity of kidney disease increases. CKD is classified into five stages, increasing in severity^2,3^.

Patients with early onset of CKD, stage 1-2, have normal to mild levels of eGFR (60 to >90mL/min per 1.73 m^2^). Patients with stages 3a-3b have moderate levels of eGFR (45-59mL/min per 1.73m^2^). Stages 4-5 are indicative of advanced stages of kidney disease and failure and are classified by severely decreased levels of eGFR (15-29 to <15mL/min per 1.73m^2^)^4^. Globally, CKD poses a significant health challenge, affecting over 10% of the population. Although mortality has declined in patients with ESKD, CKD has been recognised as one of the leading causes of mortality worldwide^5^. Therefore, it is imperative that therapeutic and preventative measures are implemented globally.

A typical therapeutic strategy to manage CKD is Vitamin D supplementation or its activated analogues, which is specifically used as a standard of treatment in chronic kidney disease-mineral and bone disorder (CKD-MBD)^6^. Due to several physiological mechanisms, there is a strong association between CKD and Hypovitaminosis D. Vitamin D deficiency is more common in CKD patients than in the general population³[, making supplementation a key therapeutic strategy. Vitamin D deficiency is typically defined by serum 25-hydroxyvitamin D [25(OH)D] levels below 50 nmol/L (20 ng/ml), with levels above this threshold considered the primary treatment target^7^. Vitamin D plays a crucial role in maintaining bone health by regulating calcium and phosphate metabolism.

Vitamin D is typically metabolised in a two-step metabolic process. Firstly, it is acquired through diet and sunlight exposure and undergoes 25-hydroxylation in the liver, forming 25-hydroxyvitamin D [25(OH)D], which is the main circulating form of vitamin D. The 25-hydroxyvitamin D [25(OH)D] (inactive form) is converted into 1,25-dihydroxyvitamin D [1,25(OH)₂D] (active form) via the enzyme 1-α-hydroxylase which is synthesised in the kidneys^8^. Reduced kidney function and nephrotic loss in CKD impair the conversion of 25(OH)D to active 1,25(OH)₂D, causing vitamin D deficiency and hypocalcaemia. This triggers PTH release via calcium-sensing receptors, leading to secondary hyperparathyroidism (sHPT), which increases bone turnover, fragility, and vascular calcification, contributing to CKD-MBD. Despite supplementation, debate remains over the effectiveness of inactive versus active vitamin D analogues on kidney function, PTH control, and bone health.

Calcium metabolism is regulated by active vitamin D and PTH in the intestines, kidneys, and bones. PTH increases bone resorption and activates 1-α-hydroxylase, while active vitamin D enhances intestinal calcium absorption to correct hypocalcaemia^10^. Currently, there are no clear guidelines for calcium intake in dialysis or post-transplant CKD patients, and calcium management remains under-researched, underscoring the need for further study on long-term risks and intake balance in CKD care.

Vitamin D deficiency is strongly associated with CKD progression, bone mineral disorders, secondary hyperparathyroidism (sHPT), and increased mortality. However, the clinical effectiveness of vitamin D and calcium supplementation individually or combined remains uncertain. For instance, Ngai (2014), reported high rates of vitamin D deficiency and elevated PTH in pre-dialysis CKD patients, while others link deficiency to increased risks of cardiovascular death, cancer, and mental health issues^12,13^. Despite evidence suggesting that vitamin D supplementation may reduce PTH, findings are inconsistent. Zisman et al. (2014) found a non-significant reduction in PTH after supplementation. Calcium supplementation also remains controversial, with mixed results some showed improved BMD, while others linked it to vascular calcification and hypercalcemia. The lack of clear guidance on calcium intake and conflicting evidence justify the need for the current study. Therefore, the study evaluated and synthesised existing evidence on the effects of vitamin D and calcium supplementation in individuals with CKD, with a particular focus on kidney function and QoL. Specifically, the review assessed the effect of supplementation on kidney function, using indicators such as eGFR and serum creatinine levels considering both clinical outcomes and patient-reported experiences. Furthermore, the review compares the effectiveness of inactive forms of vitamin D (such as cholecalciferol and ergocalciferol) with active forms (calcitriol and alfacalcidol) in influencing kidney function and QoL as well as the role of these supplements in managing CKD-related bone mineral disorders.

## Methodology

This systematic review was conducted in accordance with the protocol of PROSPERO registered with identification number CRD42024607320. To ensure methodological rigour and replicability, the review was reported following the Preferred Reporting Items for Systematic Reviews and Meta-Analyses (PRISMA) guidelines^4^.

## Study Identification

Identification and screening of studies were conducted from October 2024 to February 2025, using PubMed, Google Scholar, Science Direct, and EBSCO. Table 1 displays the search strategy, keywords and Boolean operators used in the search.

**Table 1:**
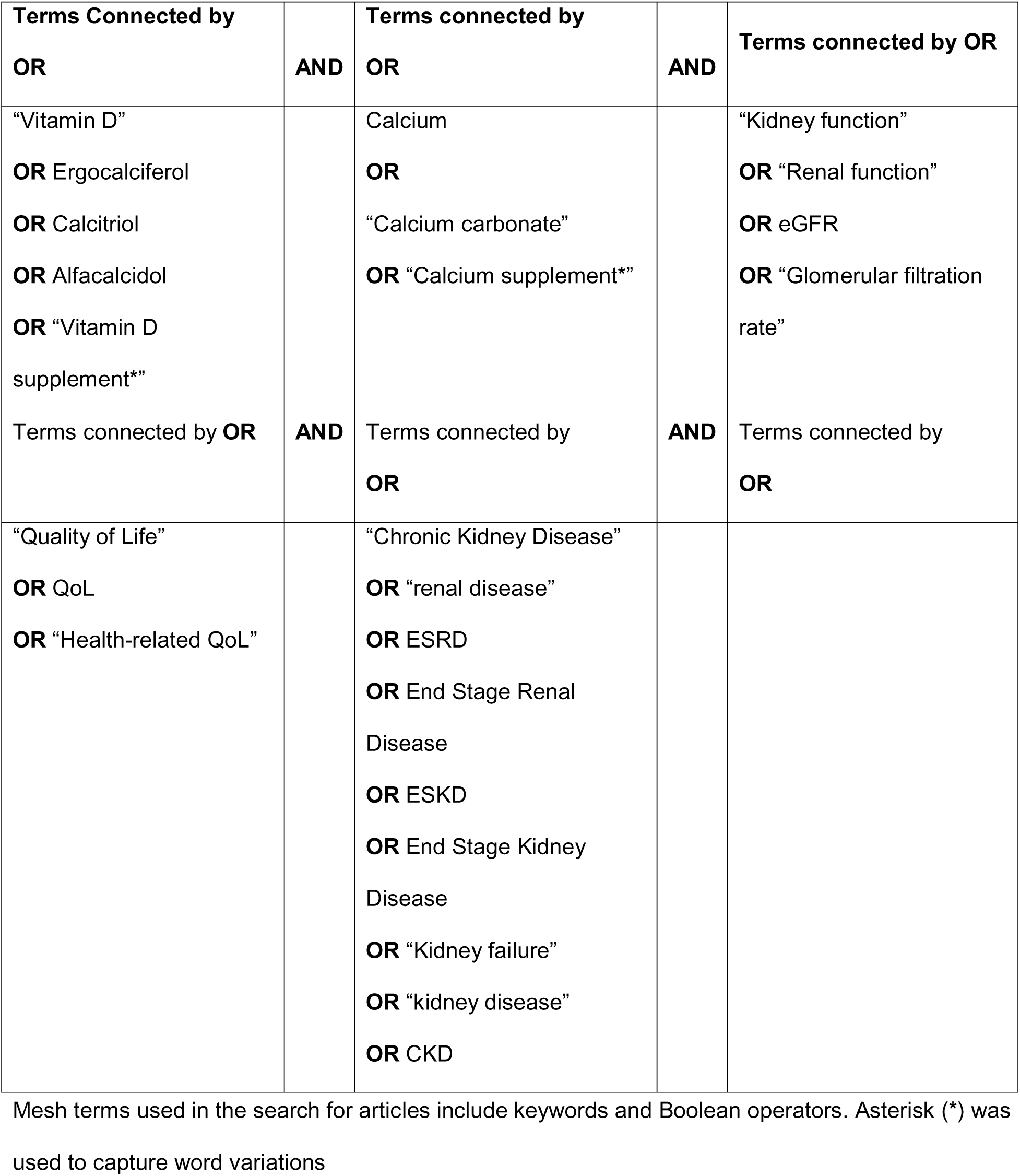
Search Strategy Illustrating Keywords and Corresponding Terms Including Mesh Terms.

## Study Selection

Inclusion criteria were randomised controlled trials, clinical trials and observational studies, which detailed the use of vitamin D and/or calcium as an intervention for adults (18 years and over) who had been clinically diagnosed with chronic kidney disease stages 1-5. studies written in English with full-text availability. The publication period was from January 2014 to December 2024. The exclusion criteria included studies on animals, children, adolescents, kidney transplant and dialysis recipients and individuals with comorbidities that significantly affect vitamin D and calcium supplementation.

Studies were initially screened based on their title and abstracts followed by full text screening. A PRISMA flow diagram was used to keep a record of the number of studies at each stage^15^. The primary outcomes assessed included kidney function measured using eGFR (mL/min/1.73m²) and serum creatinine levels (µmol/L), quality of life assessed through QoL questionnaires, bone mineral disorders (BMD) associated with CKD measured through bone turnover markers, fracture rates, serum calcium and phosphorus, and parathyroid hormone (PTH) levels.

## Data extraction and analysis

Key information from the included studies was extracted using a Microsoft Excel spreadsheet, capturing details such as authorship, year and country of publication, study design, sample size, population characteristics (including age, sex, and CKD stage), intervention specifics (type, dosage, duration, and frequency), and reported outcomes. Study quality was assessed using the Critical Appraisal Skills Programme (CASP)^16^, a widely used tool for evaluating systematic review studies. For randomised controlled trials (RCTs), risk of bias was visually presented using the Robvis tool.

Both qualitative and quantitative methods were employed in synthesising the data. A meta-analysis was conducted to examine the impact of vitamin D supplementation on eGFR, with findings displayed in a forest plot. Mean differences were calculated, and statistical heterogeneity was assessed using the Cochrane Q test and the I² statistic. As no heterogeneity was found, a fixed-effects model was used. A second meta-analysis evaluated the effect of vitamin D on PTH levels. In this case, statistical heterogeneity was detected, prompting the use of a random-effects model. Forest plots were used to present both sets of results.

To compare inactive (ergocalciferol/cholecalciferol) and active (calcitriol/alfacalcidol) forms of vitamin D, a paired sample t-test was conducted. All quantitative analyses were performed using the ‘metafor’ package in R (version 4.4.3). A narrative synthesis was applied to outcomes related to QoL and BMD, due to the heterogeneity of the data which made meta-analysis impossible. Additionally, because of limited data, calcium supplementation effects could not be meaningfully analysed.

## Results

A total of 86 records were identified through electronic database searches. Prior to screening, 14 duplicate studies were removed along with three additional studies that did not meet the eligibility criteria, leaving 69 studies to proceed to the initial screening stage. A further 28 studies were excluded due to irrelevant outcomes or inappropriate study design. After full text review, 26 studies were further excluded leaving 15 studies that were included in the final systematic review. The PRISMA flow diagram (Figure 1) provides a detailed overview of the review process.

**Fig. 1:**
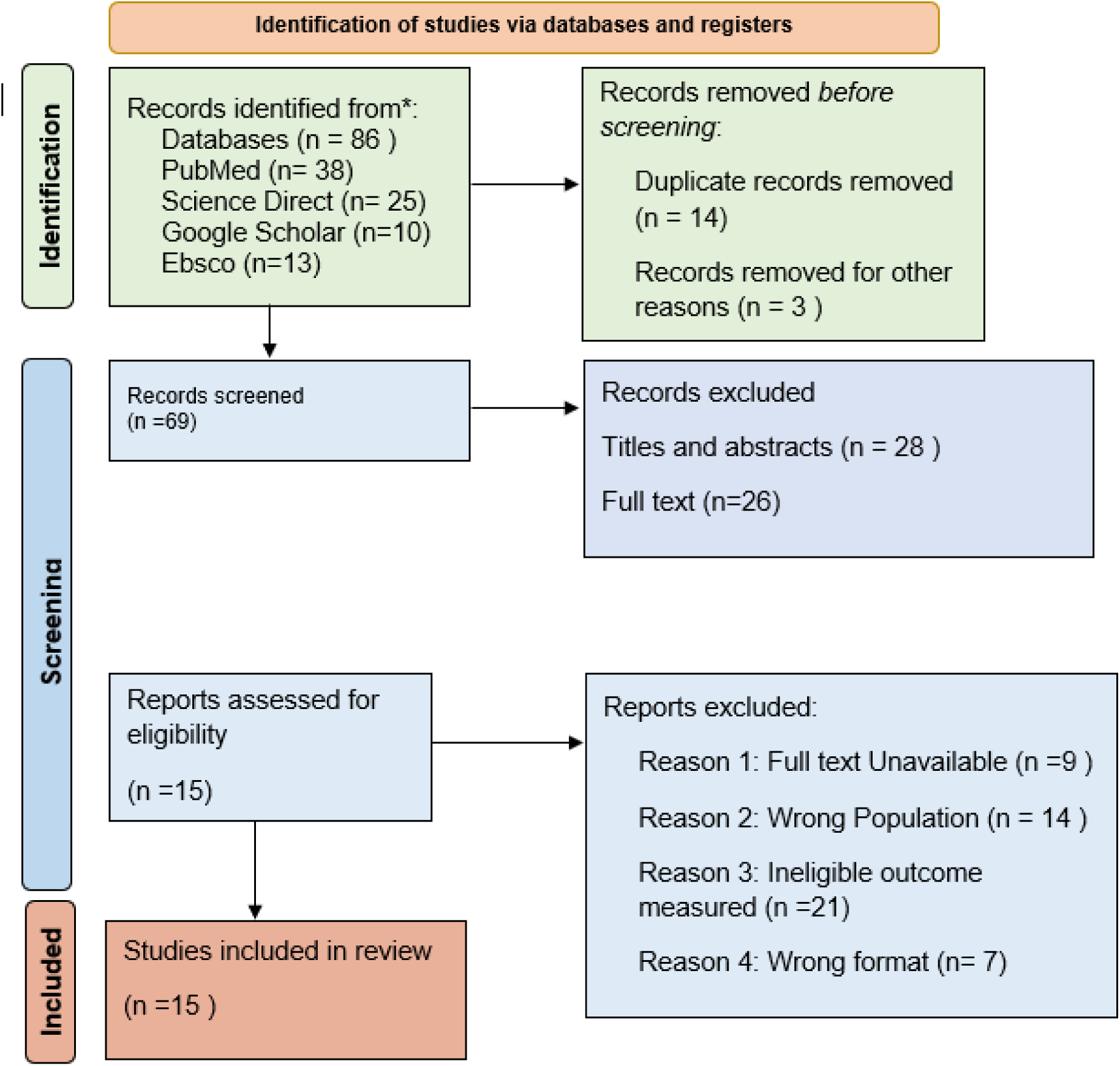
PRISMA flow diagram illustrating the study selection process for the systematic review.

## Study and participant characteristics

The 15 eligible included studies had a total of 3,354 CKD patients across ten countries. Ten of the studies were conducted in Western countries and five were of Asian origin. The majority of studies (n= 14) were randomised controlled trials (RCTs), while one study was an observational study. Most of the studies assessed vitamin D supplementation, whereas only one study investigated calcium supplementation, highlighting a gap in research on the role of calcium in CKD management. The primary outcomes investigated were kidney function, PTH levels, bone mineral metabolism and QoL. Table 2 summarizes the characteristics of the included studies.

## Risk of Bias Assessment

Quality of the studies was assessed using the CASP tool^16^. Seventy percent of the RCTs had a low risk of bias, while 30% showed some concerns. Only one study presented a high risk of bias due to selection of the reported result. The observational had a low risk of bias. The Robvis tool was used to graphically present these findings these findings in Figure 2.

**Fig. 2:**
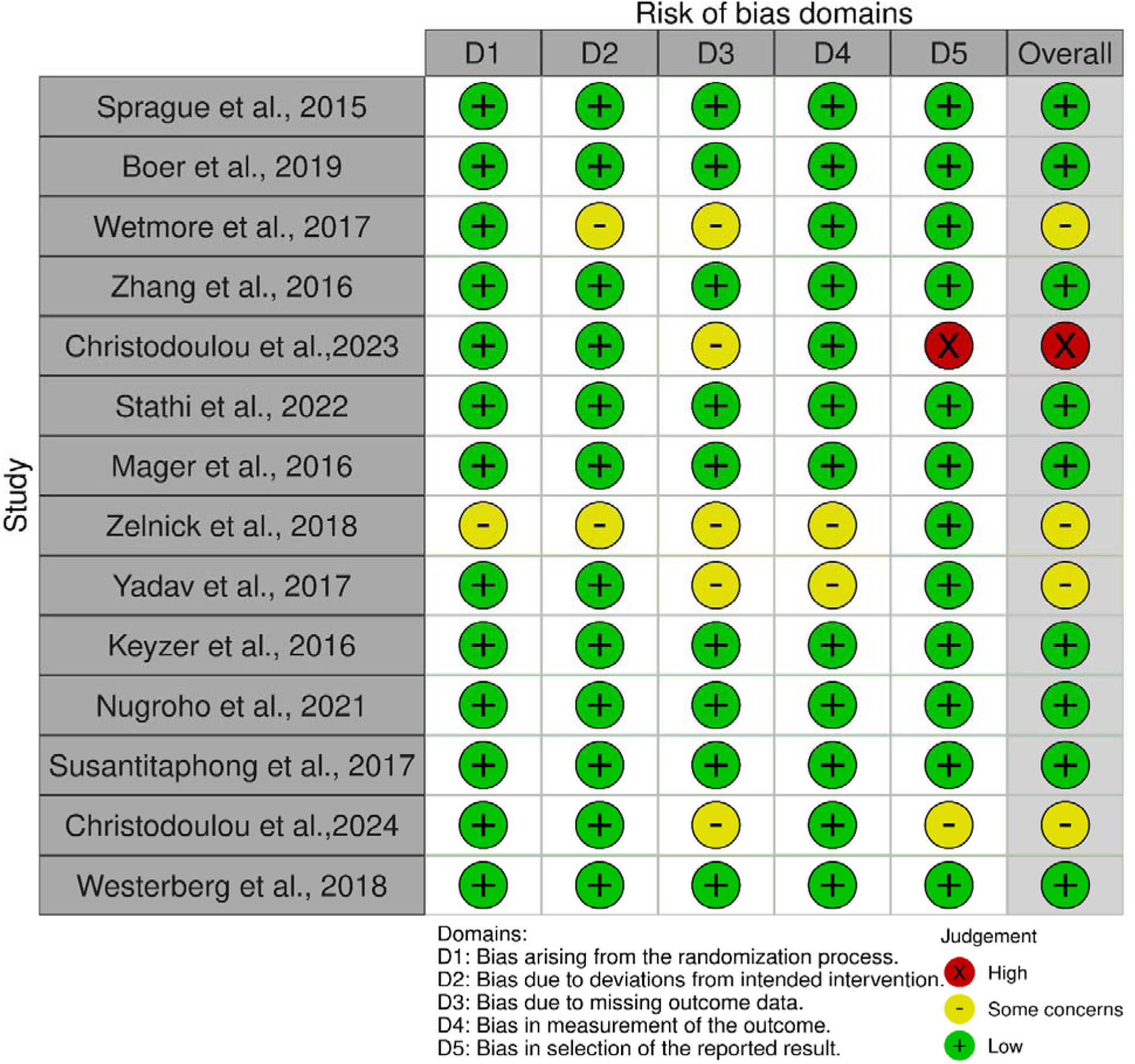
Risk of Bias assessment for RCTs. The risk of bias assessment for RCTs was categorised in five domains: D1, D2, D3, D4 and D5 all represented by an overall verdict for each study. Among the included studies, ten have low risk of bias, three studies have some concerns while two studies have high risk of bias.

## Impact of Vitamin D Supplementation on eGFR

A meta-analysis was used to assessed the impact of all types of vitamin D supplementation on eGFR in CKD patients in four studies with 451 participants^17–20^. The analysis was conducted using a fixed-effects model, with the heterogeneity assessed using the Cochrane Q statistic and I² test. The pooled results show that vitamin D supplementation had no significant impact on eGFR levels (MD=0.84mL/min/1.73m², 95% CI: –0.47, 2.15, p=0.2070). The I² statistic for heterogeneity was 0%, indicating that there was no heterogeneity among the studies. Figure 3 presents the forest plot for the meta-analysis.

**Fig. 3:**
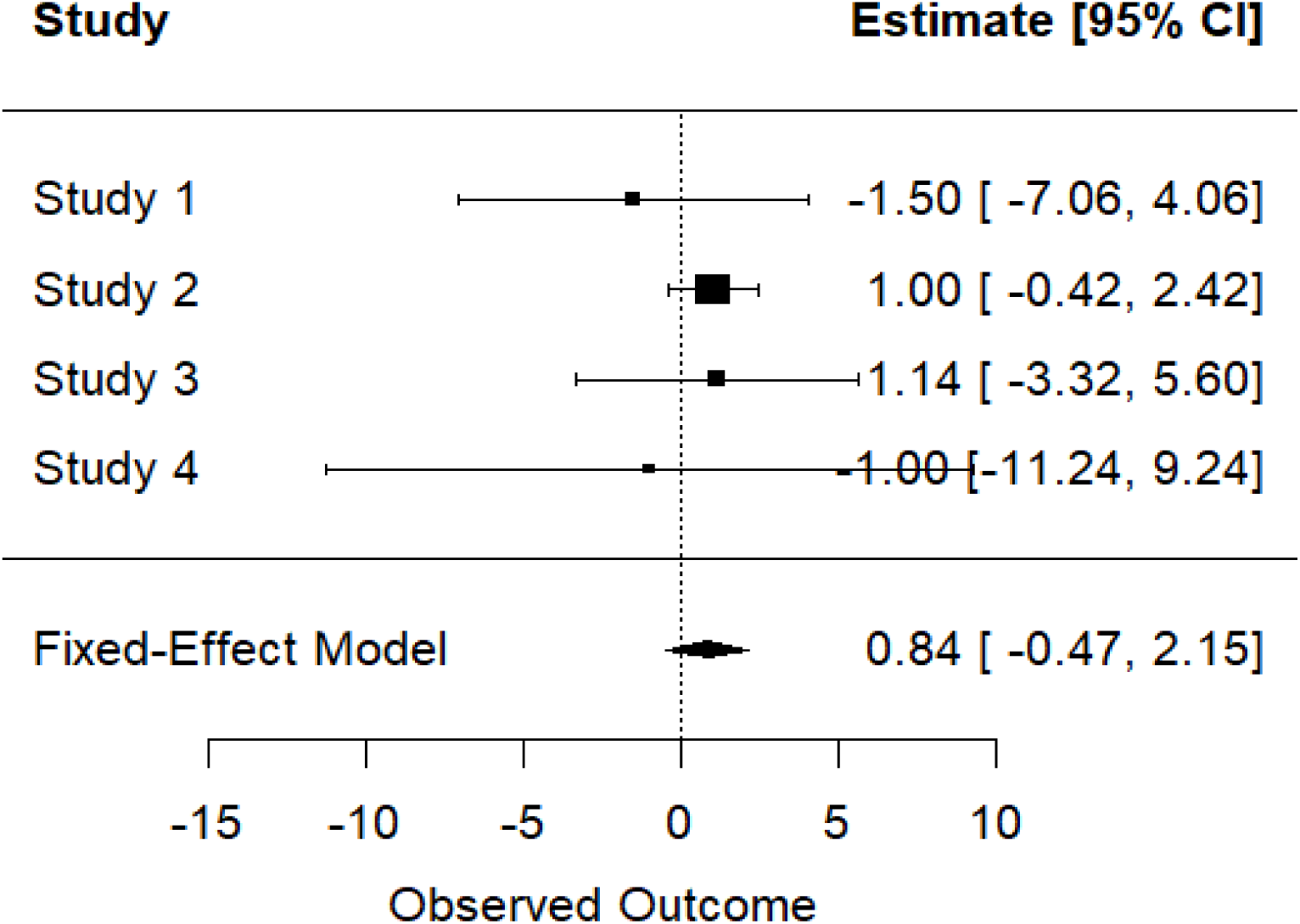
Forest plot showing the effect of vitamin D supplementation on eGFR across the included studies. Each horizontal line represents the 95% confidence interval (Cl), with he the square size reflecting the study weight in the meta-analysis. The pooled effect estimate, calculated using a fixed-effects model, is represented by the diamond at the bottom.

## Comparison of Inactive vs. Active vitamin supplementation on Serum Creatinine

A paired t-test was conducted in which the mean serum creatinine for inactive vitamin D was 11008.22 µmol/L, while for active vitamin D, it was 9607.62 µmol/L. The was no significant difference between the two groups (t= –0.093, p=0.931, 95% CI –63210.25 to 60409.05). This finding is graphically presents in a box plot in Figure 4.

**Fig. 4:**
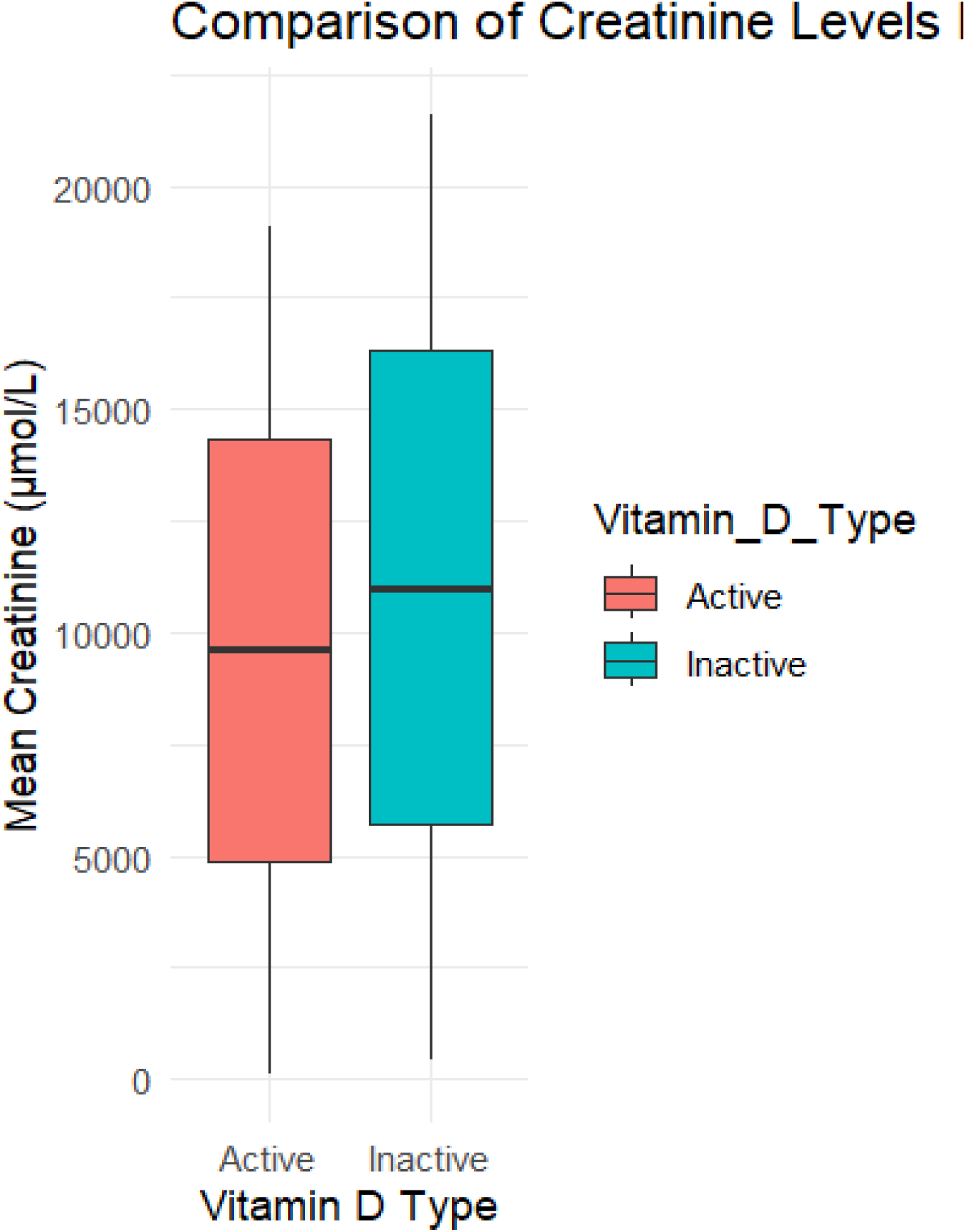
Box plot comparing the mean creatinine levels (µmol/L) between active and inactive vitamin D supplementation groups. The median creatinine levels are represented by the bold horizontal lines within each box. The interquartile range (IQR) is shown by the boxes.

## Effect of Vitamin D Supplementation on PTH Levels

A meta-analysis including a total of six studies with 522 participants assessed the effect of all types of vitamin D supplementation on PTH levels^17,21–25^. The analysis was conducted using a random-effects model, with heterogeneity assessed using the Cochrane Q statistic and I² test. The pooled results show that vitamin D supplementation did not have a statistically significant impact on PTH levels (MD=-6.76mL/min/1.73m², 95% CI –34.92 to 1.95, p= 0.005). A high heterogeneity was observed (I² = 70.87%) among the studies. Figure 5 presents the forest plot for the meta-analysis.

**Fig. 5:**
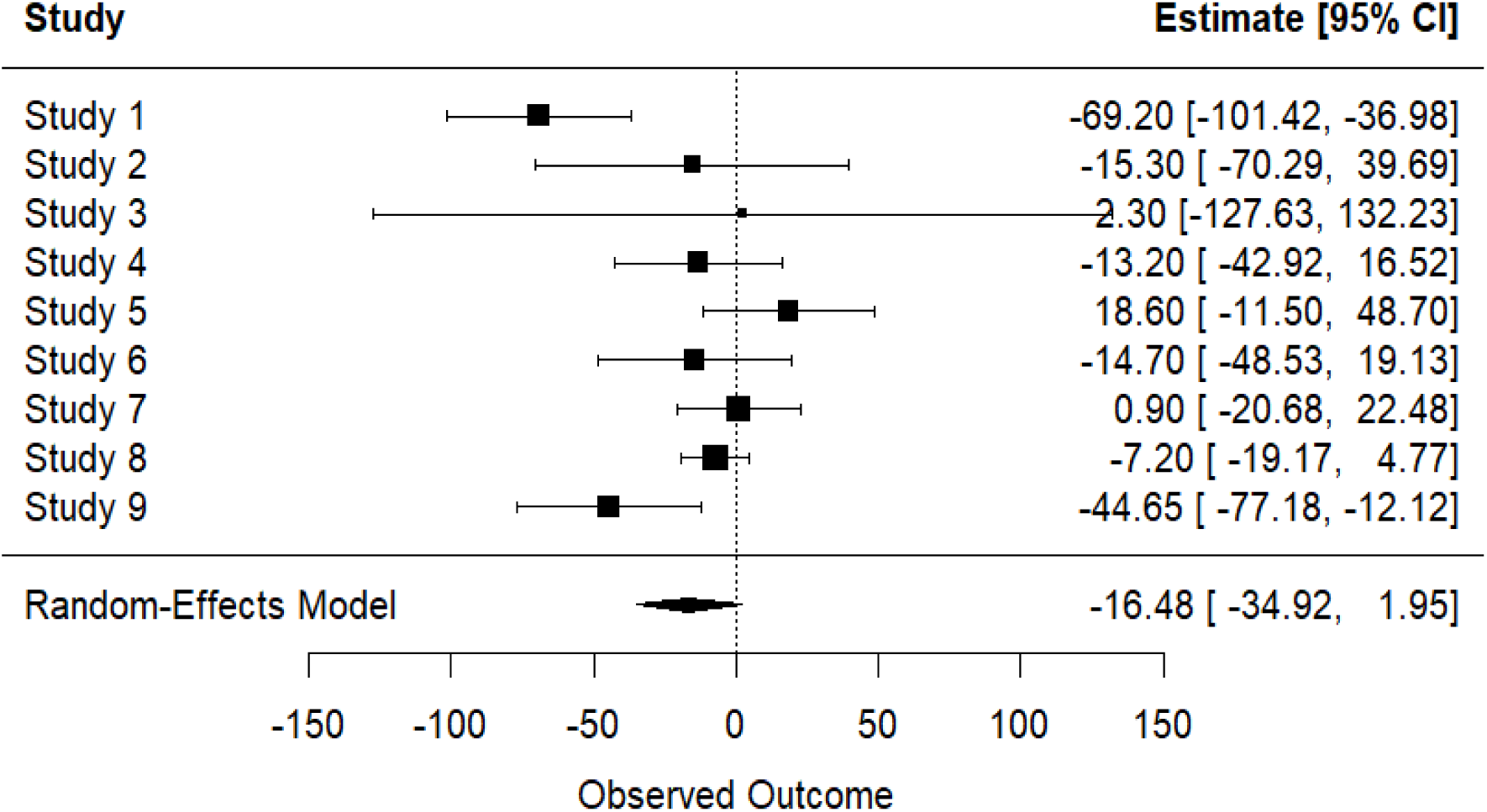
Forest plot showing the effect of vitamin D supplementation on PTH level across the included studies. Each horizontal line represents the 95% confidence interval (Cl), with the square size reflecting the study weight in the meta-analysis. The pooled effect estimate, calculated using a fixed-effects model, is represented by the diamond at the bottom.

## Influence of Vitamin D supplementation on QoL in CKD Patients

A narrative synthesis of three studies assessed the impact of vitamin D supplementation on QoL in CKD patients. Although fatigue, a common CKD symptom, has been linked to vitamin D deficiency^26^, high-dose cholecalciferol for 12 weeks showed no significant effect on fatigue levels^25^. Self-reported fatigue scores showed no improvement: physical fatigue (p=0.11), mental fatigue (p=0.25), Visual Analog (p=0.96), and hand grip strength (p=0.98). Vitamin D deficiency has also been associated with depression and anxiety^27^, but this review found no significant changes in mental health-related QoL. Sprague et al.^17^ reported no difference in Beck Depression Inventory-II scores after 6 weeks, and Mager et al.^28^ observed no mental health QoL improvements after 6 months of supplementation.

## Effect of vitamin D supplementation on Bone Mineral Density (BMD) in CKD patients

A narrative synthesis of 10 studies explored the impact of vitamin D supplementation on bone mineral density (BMD) in CKD patients, but results were inconsistent. Christodoulou et al.^18^ found no significant changes in hip or femoral neck BMD (p>0.05), though CTX-1 and iPTH decreased, while FGF-23 and BAP increased. Similarly, no significant differences in total body, spine, or hip BMD (p>0.05), with no improvements in bone metabolism markers (OC, NTx, FGF-23)^28^. Westerberg et al.^25^ also observed no BMD improvements or reduction in fracture risk following high-dose vitamin D. In contrast, Christodoulou et al.^24^ noted a slight decline in hip BMD after supplementation, despite reductions in CTX-1 and increases in BAP and PINP indicating possible changes in bone metabolism not directly reflected in BMD outcomes.

## Discussion

The current study investigated the impact of vitamin D supplementation on eGFR, PTH levels, QoL and BMD in CKD patients. Findings showed that vitamin D supplementation had no significant effect on eGFR or QoL. Although PTH levels declined after supplementation, meta-analysis results were not statistically significant. BMD outcomes also remained inconsistent. These findings align with previous reviews reporting that vitamin D does not improve eGFR albeit no harmful effects on kidney function^29,30^. The absence of eGFR improvement may relate to underlying CKD pathophysiology. Vitamin D plays a more critical role in regulating calcium and phosphate metabolism than in affecting kidney filtration. It enhances intestinal calcium absorption and helps maintain calcium balance through its interaction with PTH^31^, suggesting its primary influence lies in mineral regulation rather than eGFR. The comparison of inactive and active vitamin D supplementation further supports this conclusion. Serum creatinine levels were measured, and it was found that there were no significant differences between the two forms of vitamin D (p= 0.930). This suggests that neither form of vitamin D has a direct impact on kidney function. This outcome is expected, given that the conversion of inactive vitamin D (25-hydroxyvitamin D) to active vitamin D (1,25-dihydroxyvitamin D) occurs primarily in the kidneys^32^. In CKD patients, the loss of functional nephrons limits this conversion, potentially reducing any renal benefits of supplementation. However, variability in creatinine levels could be associated with differences in baseline eGFR and comorbidities rather than with vitamin D supplementation itself. Future studies may require the separation of CKD stages to determine how different subgroups respond differently to treatment.

Although some studies reported that vitamin D supplementation reduces PTH levels, this review and meta-analysis found no significant decline in PTH concentrations (MD = –6.76 pg/mL, p = 0.005). Kandula *et al.*^34^ found contrasting results, with an associated decline in PTH levels. It was found that the most significant reduction in PTH levels occurred in dialysis patients, suggesting that vitamin D supplementation may be more effective in late-stage CKD. However, Christodoulou, Aspray and Schoenmakers^35^ found similar inconsistencies, aligning with the findings of this review. They found that vitamin D supplementation had a non-significant suppressive effect on PTH concentrations, and there were inconsistencies between studies. The high heterogeneity (I² = 70.82%) may be due to differences in CKD stage, vitamin D dosage, and study duration. Additionally, PTH regulation involves multiple factors, including calcium and phosphate homeostasis. Since calcium plays a critical role in suppressing PTH, vitamin D alone may not be sufficient. This suggest a need to explore combination therapy with calcium and phosphate supplementation in future studies to assess whether a combined approach better manages secondary hyperparathyroidism (sHPT) in CKD patients.

Chronic kidney disease mineral bone disorder (CKD-MBD) is a condition characterised by disruptions in the calcium and phosphate balance as well as PTH and FGF-23 metabolism, which in turn promotes bone disorders^36^. The inconsistent results on BMD outcomes indicate that supplementation of vitamin D provides no meaningful improvements. Studies found that supplementation did not insignificantly improvem of total body, spine, hip and femoral neck BMD^18,28^. Christodoulou *et al.*^24^ and Christodoulou *et al.*^18^ reported a reduction in the bone turnover marker CTX-1, suggesting that bone reabsorption was reduced post-supplementation. In addition to this, an influx of bone turnover markers, BAP and PINP, suggested that there was increased bone formation, which could be a compensatory mechanism to counteract previous bone loss. Additionally, Christodoulou *et al.* ^18^ reported an increase in the hormone FGF-23; however, his later study gave contrasting results. These findings suggest that vitamin D may influence bone turnover, but is not able to directly impact BMD outcomes. Therefore, despite expectations that vitamin D supplementation would improve bone health in CKD patients by regulating calcium-phosphate metabolism, these findings suggest that vitamin D alone would not be sufficient to prevent bone loss in CKD patients. However, the study duration of these trials were typically short-term, whereas BMD changes usually take a minimum of 3 years to present adequate data ^36^. This suggests that there is potential for vitamin D supplementation to provide significant results, yet there are limited long-term studies available to convey this.

Regarding QoL, there are very limited studies on the effect of vitamin D supplementation on QoL in CKD patients; however, in previous studies, it has been found that vitamin D deficiency was significantly associated with lower scores in QoL metrics. My findings found no significant results, which is consistent with previous research^37^. Oh *et al.*^37^ found that in QoL metrics, the vitamin D deficiency group had significantly lower scores compared to those without vitamin D deficiency. The included studies in this analysis used varied QoL measurement tools, making it difficult to compare findings. Self-management interventions, such as nutritional intervention, counselling and exercise, may be more effective at improving QoL in patients with CKD^38^. Future studies could potentially investigate the combined treatment of vitamin D supplementation with self-management strategies.

## Limitations

This systematic review evaluated the impact of vitamin D and calcium supplementation in CKD patients; however, there are several limitations that should be noted. Firstly, a vast amount of heterogeneity was observed across the included studies. The variability in CKD stages, dosage and type of supplementation, duration of treatment, and outcome measures, may have contributed to the inconsistent results, and limited the ability to make clear comparisons. Many studies did not separate results into subgroups, such as CKD stage, age, or baseline vitamin D levels. This makes it difficult to identify which patients may have benefited most from supplementation. Instead, results were often combined into a single outcome, potentially withholding important differences between subgroups. Another key limitation is the variation in study duration. The majority of the included studies were largely short-term, ranging from six weeks to one year. This could have likely influenced the findings, particularly for outcomes like BMD and QoL, which often require long-term observations to detect meaningful changes. Small ^26^, highlights that changes in BMD usually take at least three years to present with adequate data, suggesting the studies included may not have been long enough to capture the impact of supplementation.

With regards to QoL, the limited number of studies and inconsistency in measurement tools, further restricted analysis. The differences in questionnaires and scoring methods used, made it difficult to compare outcomes as well as perform a meta-analysis. This lack of standardisation reduced the strength of conclusions drawn about QoL improvements. Lastly, despite this systematic review looking into both vitamin D supplementation and calcium supplementation, meaningful conclusions were not able to be drawn about calcium effects. The number of eligible studies on calcium alone was extremely limited, with only one meeting the inclusion criteria of this review. This highlights a significant gap in the literature, of which should be addressed in future research.

## Recommendations for Future Research

To strengthen future evidence, several improvements are needed in the design and scope of studies on vitamin D and calcium supplementation in CKD. Most existing research has been short-term (under one year), limiting insight into outcomes like BMD and QoL, which typically require longer periods to show meaningful changes. Future studies should therefore prioritise long-term investigations.

The limited research on calcium supplementation especially studies examining its independent effects makes it difficult to draw tangible conclusions. Future work should evaluate calcium only as well as in combination with vitamin D. Subgroup analyses would also help clarify which patient populations benefit most from supplementation. Standardising outcome measures, particularly for QoL, would improve comparability across studies. QoL remains an underexplored area in relation to these interventions and deserves greater focus. In addition, future research should monitor long-term safety outcomes, especially concerning the potential of calcium to cause hypercalcaemia and vascular calcification. Including patient-reported outcomes could further highlight the real-world impact of these interventions on daily living.

## Conclusion

This systematic review assessed the effects of vitamin D and calcium supplementation on kidney function and QoL in CKD patients. Findings showed no significant impact of vitamin D on eGFR or QoL. Although PTH levels declined post-supplementation, the change was not statistically significant. Bone turnover markers suggested shifts in bone metabolism, but these did not reflect in BMD outcomes. Due to limited data, no conclusions could be drawn regarding the effects of calcium supplementation on kidney function or QoL. The review highlights the need for a more integrated approach to managing CKD-related outcomes, including bone and mineral disorders. Future studies should explore combined supplementation with vitamin D, calcium, and phosphate, while also accounting for CKD stage and baseline vitamin D status. Also, it is appropriate to prioritise long-term randomised controlled trials and evaluate combination therapies to determine whether sustained supplementation offers meaningful benefits for kidney function, QoL, and bone health in CKD patients in future studies.

## Data Availability

All data produced in the present work are contained in the manuscript

---

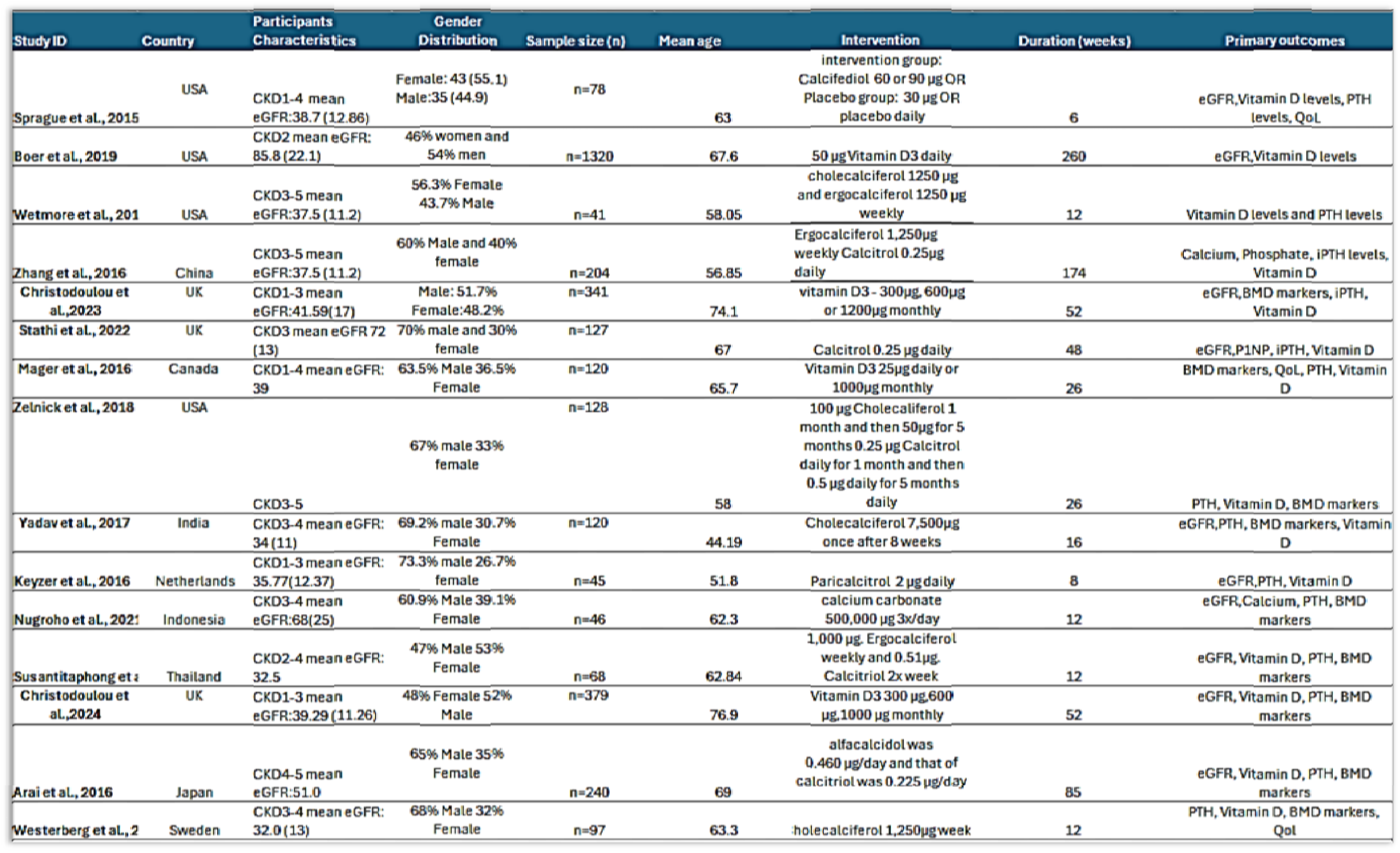

## Notes

### Competing Interest Statement

The authors have declared no competing interest.

### Funding Statement

This study did not receive any funding

